# Water Supply Continuity, Frequency, and Health Gains: Ten-year Evidence from Hubli-Dharwad, India

**DOI:** 10.64898/2026.07.23.26358804

**Authors:** Ayse Ercumen, Narayana Billava, Zachary Burt, Sharada Prasad, Nayanatara Nayak, Emily Kumpel

**Affiliations:** North Carolina State University, Raleigh, NC, USA; Center for Multidisciplinary Development Research, Dharwad, India; California Department of Water Resources, Sacramento, CA, USA; Infosys, Bengaluru, India; University of Massachusetts, Amherst, MA, USA

## Abstract

Intermittent water supplies (IWS) serve >1 billion people globally and can transmit waterborne infections. How often and for how long supply is delivered varies between and within IWS systems. The UN Sustainable Development Goals target having water “available when needed” for ≥12 hours/day or ≥4 days/week but there are scarce data on how supply frequency/duration within IWS affect health outcomes. We conducted a matched study in Hubli-Dharwad, India, a city served partially by continuous water supply (CWS) since 2007 and partially by IWS. We enrolled 2220 CWS and 2218 IWS households matched on socioeconomics and sanitation. We compared diarrhea prevalence in children <5 years and typhoid fever incidence between households with different water supply characteristics using generalized linear models with robust standard errors and adjusting for socio-demographics and sanitation. Among IWS households, the median supply frequency was every 8 days (interquartile range [IQR]=7-8), and the median supply duration was 4 hours (IQR=3-5). IWS households had 36% higher prevalence of child diarrhea (prevalence ratio [PR]=1.36, 1.00-1.84) and 78% more typhoid fever cases (cumulative incidence ratio [CIR]=1.78, 1.05-3.02) than CWS households. IWS households meeting the UN criterion and those in the top quintiles of supply frequency and duration (receiving water once every 1-6 days or for 7-24 hours per supply cycle) had similar health outcomes as CWS households. IWS households below the UN criterion had higher diarrhea prevalence (PR=1.41, 1.03-1.93) and more typhoid fever cases (CIR=1.93, 1.15-3.22) than CWS households, as well as more typhoid fever cases than IWS households meeting the criterion (CIR=3.66, 1.37-9.79). IWS households in the bottom quintiles of supply frequency and duration (receiving water once every 9-15 days or for ≤3 hours per supply cycle) had 45-72% higher child diarrhea prevalence and twice as many typhoid fever cases than CWS households (p-values<0.05). These findings support global efforts to implement CWS. Our results also highlight that increasing supply frequency and duration in IWS systems in the interim can deliver health benefits, and the UN criterion of having “water available when needed” improves health.

## Introduction

Over 1 billion people worldwide are served by intermittently operated piped water systems where water is delivered at intervals [1]. Intermittent delivery deteriorates water quality in the pipes through intrusion of contaminants in the absence of continuous pressure in the pipes [2]. It also necessitates water storage in the home between supply cycles, which can introduce contamination at the point of use [3,4]. Households with intermittent supply may rely on additional non-piped water sources with inferior water quality and may also be limited in the quantity of water available for personal and domestic hygiene. Consequently, intermittent water supply has been associated with increased risk of diarrheal disease [5–7] and outbreaks of waterborne diseases such as typhoid fever and cholera [8–11]. Interruptions in water supply have also been associated with respiratory infections [12,13] and stress [14].

While health effects from intermittent piped water supplies have been studied compared to continuous supplies [15], less attention has been given to heterogeneity in the interval (days between supply) and duration (hours of supply) within intermittent systems. Supply intervals can vary widely both between and within intermittent systems, ranging from once a day to once a every 7+ days and with longer gaps between supply cycles in arid regions, during dry seasons and in peri-urban areas or informal settlements [16–18]. Similarly, supply duration and pressure can vary based on water availability as well as topography, with elevated areas or the peripheries of the distribution network receiving shorter supply duration and/or inconsistent pressure [19]. The United Nations Sustainable Development Goals recognize water availability as one of the four pillars of a safely managed drinking water supply [20]. The WHO/UNICEF Joint Monitoring Program tracks water availability using the indicator of “available when needed” which they define as households reporting their water is “sufficient” or available “most of the time” (at least 12 hours/day or 4 days/week) [21]. As increasing water scarcity and variability forces more utilities around the world to provide intermittent supplies, there is no robust evidence to guide decisions on optimal supply intervals or duration for intermittent systems.

We previously conducted a matched cohort study in Hubli-Dharwad, India, to evaluate the water quality and health impacts of a pilot program that provided continuous water supply (widely known in India as “24×7” supply) to 10% of the city’s population while the rest continued to receive water intermittently [22]. This study was conducted approximately three years after the upgrade and showed that areas with continuous supply had lower prevalence of bloody diarrhea among children <5 years living in low-income households and lower incidence of typhoid fever than areas with intermittent supply; there was no difference in the prevalence of diarrhea or in weight-for-age z-scores for children <5 years between intermittent vs. continuous supply areas [23]. We also found that >90% of households in continuous supply areas continued to store drinking water [24]. Tap water and stored water quality were both improved under continuous supply but contamination in storage attenuated the water quality gains between the continuous and intermittent supply areas [25]. A recent analysis of these data found that intermittent supplies that were delivered predictably (at a predetermined day/hour without delay) delivered health gains similar to continuous water supply while unpredictably delivered intermittent supplies were associated with health risks [26], indicating there are nuances within intermittent systems that influence health outcomes.

Since our previous evaluation, Hubli-Dharwad has continued to provide continuous water supply to the original pilot areas, as well as expanded efforts to improve supply continuity in additional areas of the city, but the gradual implementation has resulted in heterogeneous water access across the city. Within intermittent supply areas, a range of water delivery intervals and durations are implemented, and within continuous supply areas, sporadic interruptions are reported. We conducted a follow-up study approximately 12 years after the initiation of continuous water supply to assess the long-term impact of continuous vs. intermittent water supply on the risk of diarrhea and typhoid fever in low-income households in Hubli-Dharwad. We also aimed to assess associations between these health outcomes and the length of intervals between supply cycles and duration of water delivery per supply cycle within intermittent supply areas. We focused on lower-income households because they are less likely to own large overhead storage tanks that mitigate the effects of intermittencies.

## Methods

### Delivery of continuous water supply

Hubli-Dharwad has 67 administrative areas (wards). In 2007-2008, 8 wards were upgraded to pilot delivery of continuous water supply (CWS). These wards were selected by the local water authority to have hydraulically isolatable pipe segments and to be socioeconomically representative of the population of Hubli-Dharwad [27]. Since the pilot upgrade, additional wards were upgraded to receive CWS. At the onset of the current study (Oct 2018), the city estimated that 19 wards were served fully by CWS, 20 wards were partially served by CWS, and 28 wards remained on intermittent water supply (IWS).

### Participant selection

We conducted an initial screening survey (n=4000) in all 67 wards to estimate the percentage of the ward population served by continuous vs. intermittent supply (unpublished data). Using the screening data, we identified 16 wards where >50% of the population received CWS; all of these wards were selected for participation in the current study as treatment wards. Among the remaining wards where <50% of the population received CWS, we selected 16 control wards using multivariate matching with a genetic matching algorithm [28]. Using available data from a representative city-wide survey of 15,400 Hubli-Dharwad residents conducted in 2006 before the water system upgrades [29], we identified 16 control wards that matched the treatment wards on socioeconomic indicators, sanitation conditions and pre-upgrade water service.

In each selected ward, we systematically enrolled 100 to 150 households for a total targeted sample size of 4000 households (2000 per study arm). We divided wards into homogeneous clusters based on existing neighborhood boundaries. In each cluster, field staff started at a pre-identified landmark (e.g., temple, mosque, bank) and systematically approached households for enrollment until sample size targets were met. Households were eligible to participate in the study if had a child <5 year and used the municipal piped supply as their primary source of drinking water. In treatment wards, households were only enrolled if they had CWS. In control wards, households were only enrolled if they had IWS. In both groups, households were excluded from enrollment if they had a concrete roof. This criterion was imposed because households with weightbearing roofs can install large-volume overhead tanks which simulate a continuous water supply by providing water to the indoor plumbing system during intermittencies. By enrolling homes with no concrete roofs, we also intended to capture lower income households at higher risk of waterborne disease.

### Data collection

Field staff conducted a structured questionnaire and spot check observations in enrolled households. We recorded the caregiver-reported 7-day prevalence of diarrhea and blood in stool among children <5 years. We defined diarrhea as ≥3 loose stools within a 24-hour period. We also recorded whether any household member had typhoid fever in the last 2 years, how many cases the household experienced during that window, and how the cases were diagnosed (by laboratory testing, diagnosed by clinician based on symptoms or self-diagnosed). We recorded the 7-day prevalence of ear infections among children <5 years as a negative control outcome that should be causally independent from water supply continuity [30]. We collected data on the household’s water supply, including the interval and duration of water delivery in intermittent supply wards, and drinking water treatment and storage practices. We also recorded sanitation and hygiene conditions, including latrine access, presence of any open drains and observed presence of water and soap for handwashing in the latrine and kitchen areas, as well as demographics (age, gender, religion, caste) and socioeconomic indicators (occupation, asset ownership, housing type, whether the households has a ration card – a government-issued card for food subsidies for low-income households).

### Ethics

The study protocol was approved by the Institutional Review Board of University of Massachusetts, Amherst (2018-5063) and with an Institutional Reliance Agreement (21033) by North Carolina State University. Participants provided verbal informed consent prior to enrollment; verbal rather than written consent was sought to allow enrolling a representative study sample regardless of literacy.

### Statistical analysis

#### Outcomes

Our primary outcome was the 7-day caregiver-reported prevalence of diarrhea in children <5 years. Diarrhea was defined as 3+ loose or watery stools in a 24-hour period. Secondary outcomes were the 7-day caregiver-reported prevalence of diarrhea or blood in stool in children <5 years, the incidence of typhoid fever in the household in the last 2 years (whether the household had any cases, number of cases). We considered the incidence of laboratory-confirmed typhoid fever as an additional outcome for a sensitivity analysis.

#### Estimation strategy

We compared outcomes between continuous vs. intermittent water supply wards. Additionally, we assessed associations between health outcomes and the interval and duration of water delivery within intermittent supply wards. We defined households as meeting the SDG criterion if they received water for at least 4 days/week or 12 hours/day and compared outcomes between household meeting vs. falling below this criterion. Additionally, we defined interval as the typical number of days between the household’s two successive supply cycles and duration as the number of hours water is typically delivered during a supply cycle. We created a composite water availability variable by dividing the supply duration by the number of days between supply cycles to estimate average availability in number of hours per day. We grouped households into quintiles of water supply interval, duration and average availability and compared outcomes between each quintile vs. continuous supply, as well as between the lower quintiles (i.e. households with more days between supply cycles and/or fewer hours of supply) vs. the top quintile (i.e. top 20% of households with the fewest days between supply cycles and/or longest hours of supply).

We used generalized linear models with a Poisson error distribution and a log link to estimate prevalence ratios (PR) and cumulative incidence ratios (CIR) for these comparisons. Models used robust standard errors at the ward level to account for correlated outcomes between households in the same ward and children in the same household. We used the number of people living in the household as the offset in the models for the number of typhoid fever cases.

We conducted unadjusted and adjusted analyses. For the comparison between continuous vs. intermittent supply wards, the unadjusted analysis relies on matching in the design phase to achieve an unbiased comparison, and the adjusted analysis is intended to control for any residual confounding. We considered the following adjustment covariates: Child age and sex, mother’s education, household size, on-site latrine access, presence of open drains, presence of soap near the food preparation and latrine areas, religion, caste, housing type, having a ration card (given to low-income households to provide food subsidies), source of income and asset-based wealth quartile constructed by principal components analysis of reported asset ownership. We expect these variables to be predictive of our health outcomes and potentially associated with water supply characteristics but not on the causal pathway between water supply and health endpoints. Binary or categorical covariates with <5% or >95% prevalence were excluded from models.

### Sample size and statistical power

Our sample size of 4000 households was designed to yield 80% power to detect 24% relative reduction in diarrhea prevalence between the two study groups with a one-sided alpha of 5%, assuming 1.4 children <5 years per household, 10% diarrhea prevalence among children in intermittent supply areas, and an intra-cluster correlation coefficient of 0.005 for households within the same ward and 0.1 for children within the same household. The assumed values for these parameters were derived from data collected in our previous matched cohort study in Hubli-Dharwad [23].

## Results

### Enrollment and household characteristics

We enrolled 4438 households (2218 with IWS, 2220 with CWS) between October 2018 and January 2019. The average household size was 6.4 (Table S1). Mothers of children <5 years were on average 26 years old, and 12% had no education. The majority (64%) of households were Hindu, and 23% belonged to a scheduled caste or tribe while 89% of households held a ration card. Most (83%) of households had access to a non-shared private latrine but less than a third had soap or detergent observed in the latrine. Approximately half (58%) of households had an open sewer drain observed adjacent to their home (on/at the edge of their property or on their street). Households with IWS vs. CWS had similar characteristics; however, households with IWS appeared somewhat less likely to have soap in the latrine area (27% vs. 36%) and kitchen area (8% vs. 14%) (Table S1).

### Water supply characteristics

Among 2218 IWS households, 5% (229) met the UN goal of water availability for at least 12 hours/day or 4 days/week. The median interval between supply cycles was 8 days (interquartile range [IQR]=7-8 days, range: 1-15 days), the median supply duration was 4 hours (IQR=3-5 hours, range=1-24 hours), and the median water availability was 0.5 hours/day (IQR=0.4-0.8 hours/day, range=0.1-20 hours/day) (Table 1). Among 2220 CWS households, half (51%, 1131) experienced an interruption in the last 2 weeks and 75% (1654) in the last 3 months; the median duration of the last interruption was 8 hours (IQR=5-24 hours) (Table 1).

**Table 1.** Water supply characteristics by study arm.

|  | Intermittent supply<br>N=2218 | Continuous supply<br>N=2220 |
| --- | --- | --- |
| Supply frequency (days), mean (SD) | 6.9 (2.3) | -- |
| Supply frequency (days), median (IQR, range) | 8 (IQR=7-8,<br>range=1-15) | -- |
| Supply duration (hours), mean (SD) | 4.6 (2.4) | -- |
| Supply duration (hours), median (IQR, range) | 4 (IQR=3-5,<br>range=1-24) | -- |
| Average supply availability (hours/day), mean (SD) | 1.1 (2.1) | -- |
| Average supply availability (hours/day), median (IQR, range) | 0.5 (IQR=0.4-0.8,<br>range: 0.1-20) | -- |
| Had intermittency in last 2 weeks, % (n) | -- | 51.0 (1131) |
| Number of intermittencies in last 2 weeks, mean (SD) | -- | 2.5 (2.4) |
| Number of intermittencies in last 2 weeks, median (IQR, range) | -- | 2 (IQR=1-3,<br>range=1-15) |
| Had intermittency in last 3 months, % (n) | -- | 74.5 (1654) |
| Number of intermittencies in last 3 months, mean (SD) | -- | 5.3 (11.5) |
| Number of intermittencies in last 3 months, median (IQR) | -- | 2 (1-5) |
| Duration of last intermittency (hours), mean (SD) | -- | 19.1 (24.6) |
| Duration of last intermittency (hours), median (IQR) | -- | 8 (5-24) |
| Tap water looks or smells dirty, % (n) |  |  |
| Always/often | 9.6 (212) | 3.7 (83) |
| Sometimes | 17.2 (382) | 18.3 (407) |
| Rarely | 5.7 (126) | 10.6 (235) |
| Never | 67.2 (1490) | 66.5 (1477) |
| Owner of drinking water source, % (n) |  |  |
| Own | 75.9 (1683) | 79.7 (1769) |
| Neighbor/landlord | 22.6 (502) | 20.0 (443) |
| Public | 1.4 (31) | 0.3 (7) |
| Household obtains water from other sources, % (n) | 10.5 (232) | 0.5 (10) |
| Household stores water for drinking, % (n) | 99.5 (2207) | 91.8 (2038) |
| Source of glass of water, % (n) |  |  |
| Storage | 93.2 (2067) | 83.1 (1845) |
| Tap | 0 | 9.6 (213) |
| Filter/dispenser/bottle | 4.5 (99) | 5.4 (120) |
| Treatment status of glass of water, % (n) |  |  |
| Not treated | 44.5 (987) | 53.2 (1182) |
| Cloth filter | 42.3 (937) | 32.3 (716) |
| Boiling | 6.1 (136) | 8.2 (182) |
| Filter | 2.2 (49) | 2.8 (63) |
| Water present for handwashing in toilet area, % (n) | 46.8 (1037) | 45.4 (1008) |
| Water present for handwashing in kitchen area, % (n) | 83.1 (1844) | 64.0 (1421) |

The vast majority (92%, n=2038) of households with CWS were observed to store water for drinking (Table 1). Obtaining water from a secondary source was reported by 10% (232) of IWS households and <1% (10) of CWS households. Across both groups, 45-53% of households reported no water treatment, 32-42% reported using a cloth filter, <10% reported boiling and <3% reported using a filter.

### Waterborne disease

Among 3172 children <5 years in CWS households, 5.5% (174) had diarrhea in the past 7 days as reported by their caregiver, and 6.0% (189) had diarrhea or blood in stool (Table 2). Among all enrolled households, 5.4% (238) reported at least one case of typhoid fever among household members in the last 2 years. Of these, 78.2% (186) reported that the case was diagnosed by laboratory testing while 19.8% (47) was diagnosed by a physician based on symptoms and 2.1% (5) was self-diagnosed. Among 2220 CWS households, 4.3% (95) had at least one case of typhoid fever among household members in the last 2 years (Table 2).

**Table 2.**
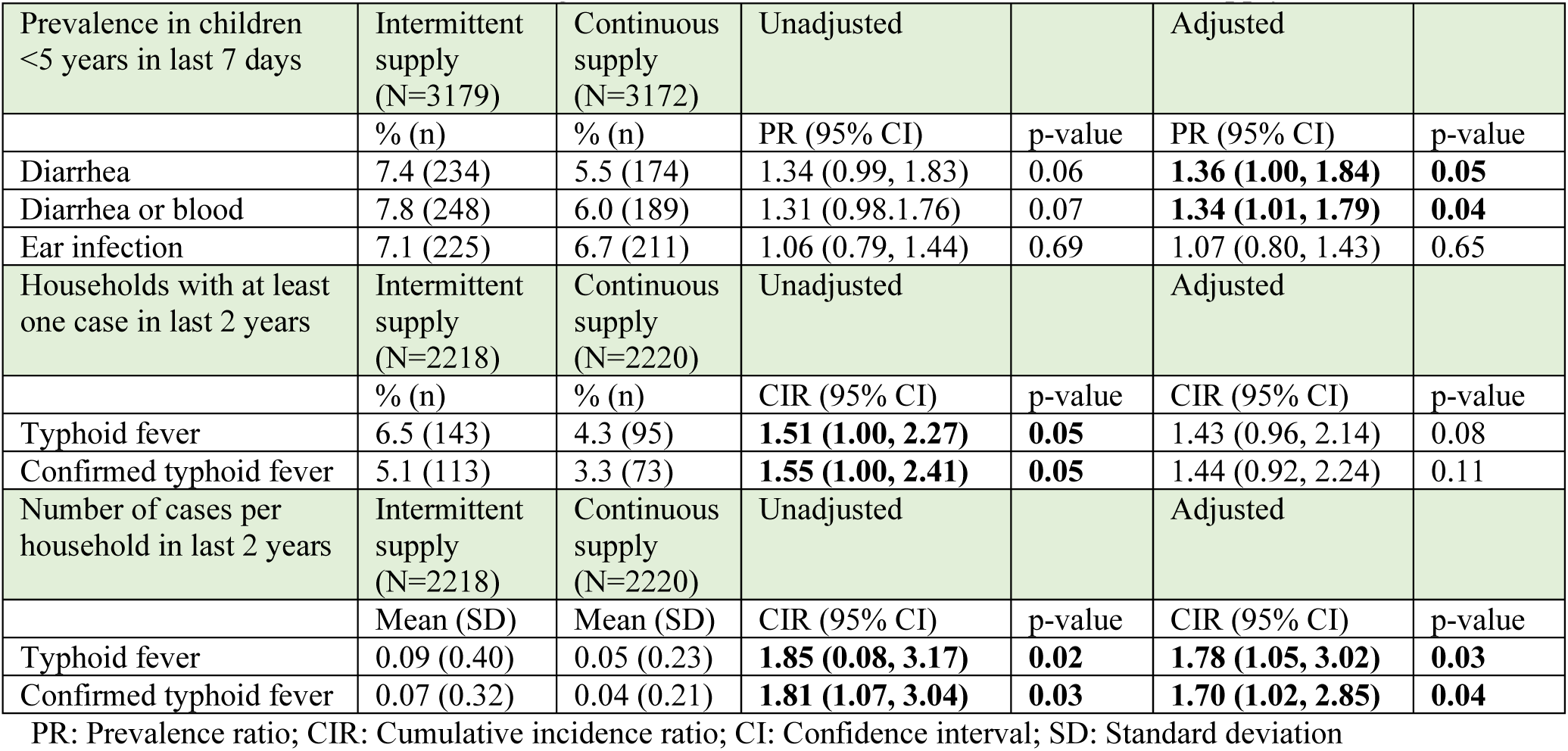
Waterborne infections among households with intermittent vs. continuous supply.

### Intermittent vs. continuous supply

In adjusted analyses controlling for potential confounders, children <5 years with IWS had 36% higher 7-day prevalence of diarrhea (PR=1.36, 1.00-1.84) than those with CWS (Fig 1, Table 2). Findings were similar for the 7-day prevalence of diarrhea or blood in stool. Households with IWS were 43% more likely to have experienced ≥1 typhoid fever case in the last 2 years (CIR=1.43, 0.96-2.14) and had 78% more typhoid fever cases (CIR=1.78, 1.05-3.02) as those with CWS (Fig 1, Table 2). Findings were similar for laboratory-confirmed typhoid fever. There was no association between IWS vs. CWS and the 7-day prevalence of ear infection among children <5 years, our negative control outcome (Table 2).

**Fig 1.**
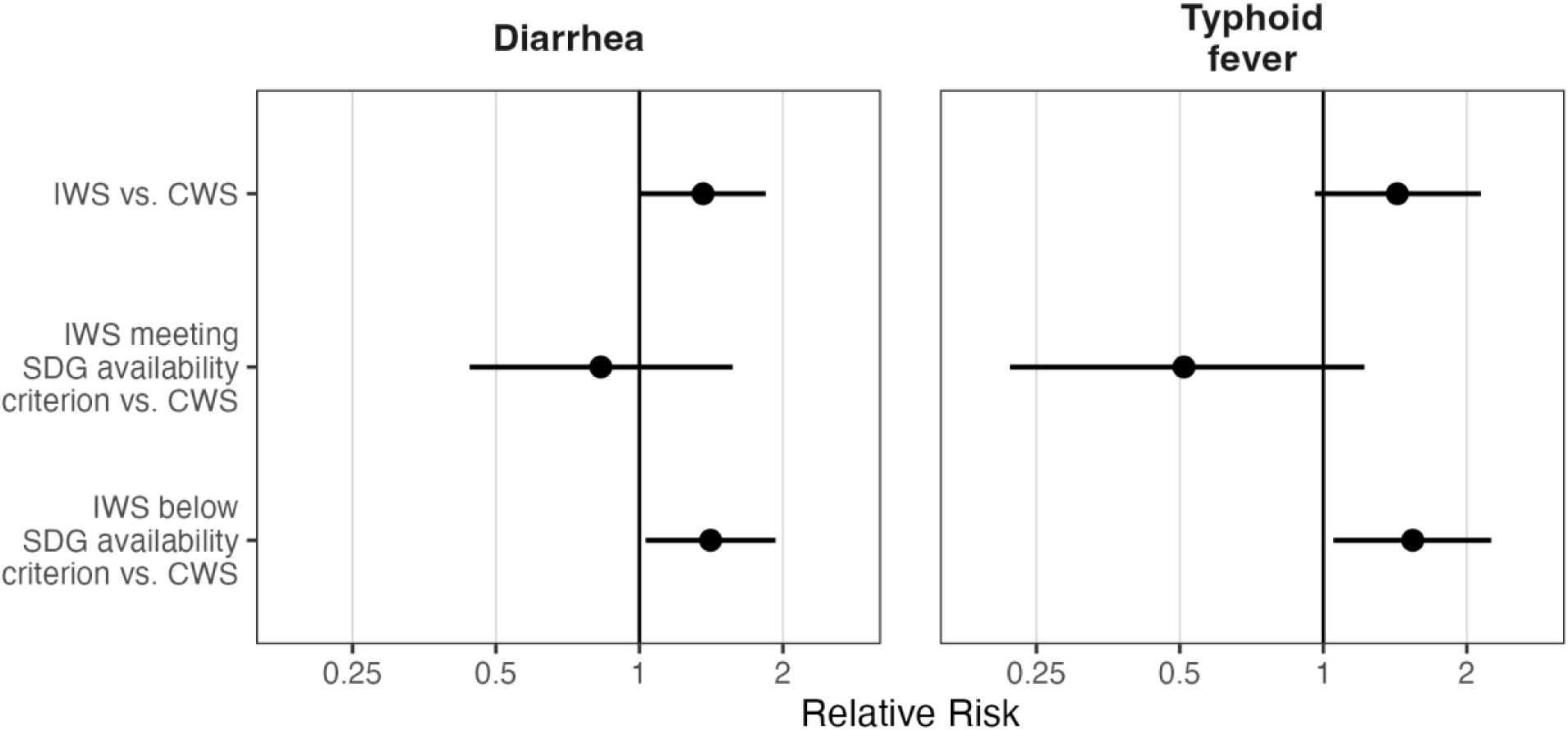
Relative risks for diarrhea prevalence in children <5 years and typhoid fever among household members for households with intermittent water supply (IWS), households whose intermittent supply meets the water availability criterion of the United Nations Sustainable Development Goals (SDGs), and households whose intermittent supply is below the water availability criterion of the SDGs, compared to households with continuous water supply (CWS). Circles denote point estimates; horizontal lines denote 95% confidence intervals.

### SDG criterion of water availability

Among IWS households, those that met the SDG criterion of having water supply for at least 4 days/week or 12 hours/day had health outcomes similar to CWS households (Fig1, Table 3). Compared to CWS households, IWS households that fell below the SDG criterion had 41% higher prevalence of child diarrhea (PR=1.41, 1.03-1.93), were 54% more likely to have ≥1 typhoid fever case in the last 2 years (CIR=1.54, 1.05-2.25) and had twice as many typhoid fever cases (CIR=1.93, 1.15-3.22) (Fig 1, Table 3). Compared to IWS household that met the SDG criterion, these households were also 3 times as likely to have ≥1 typhoid fever case (CIR=3.03, 1.37-6.66) and had more than 3 times as many typhoid fever cases (CIR=3.66, 1.37-9.79) (Table 3). Findings were similar for the prevalence of diarrhea or blood in stool and the incidence of laboratory-confirmed typhoid fever (Table 3). Meeting the SDG criterion was not associated with the prevalence of ear infections (Table 3).

**Table 3.** Waterborne infections among households above vs. below by the Sustainable Development Goal (SDG) criterion of having water “available when needed”, defined as having supply for at least 4 days/week or 12 hours/day.

| Prevalence in children <5 years in last 7 days | Intermittent supply not meeting SDG (N=2859) | aPR compared to continuous supply |  | aPR Compared to intermittent supply meeting SDG |  | Intermittent supply meeting SDG (N=320) | aPR compared to continuous supply |  |
| --- | --- | --- | --- | --- | --- | --- | --- | --- |
|  | % (n) | aPR (95% CI) | p-value | aPR (95% CI) | p-value | % (n) | aPR (95% CI) | p-value |
| Diarrhea | 7.7 (220) | **1.41 (1.03, 1.93)** | **0.03** | 1.61 (0.95, 2.75) | 0.08 | 4.4 (14) | 0.83 (0.44, 1.57) | 0.57 |
| Diarrhea or blood | 8.0 (2859) | **1.37 (1.03, 1.83)** | **0.03** | 1.39 (0.79, 2.09) | 0.31 | 5.6 (18) | 1.03 (0.58, 1.84) | 0.92 |
| Ear infection | 6.9 (197) | 1.05 (0.77, 0.42) | 0.78 | 0.81 (0.54, 1.20) | 0.29 | 8.8 (28) | 1.30 (0.87, 1.96) | 0.21 |
| Households with at least one case in last 2 years | Intermittent supply not meeting SDG (N=1989) | Compared to continuous supply |  | Compared to intermittent supply meeting SDG |  | Intermittent supply meeting SDG (N=229) | Compared to continuous supply |  |
|  | % (n) | aCIR (95% CI) | p-value | aCIR (95% CI) | p-value | % (n) | aCIR (95% CI) | p-value |
| Typhoid fever | 6.9 (138) | **1.54 (1.05, 2.25)** | **0.03** | **3.03 (1.37, 6.66)** | **0.01** | 2.2 (229) | 0.51 (0.22, 1.22) | 0.13 |
| Confirmed typhoid fever | 5.5 (109) | **1.55 (1.02, 2.37)** | **0.04** | **3.15 (1.68, 5.92)** | **<0.005** | 1.8 (4) | **0.50 (0.25, 1.00)** | **0.05** |
| Number of cases per household in last 2 years | Intermittent supply not meeting SDG (N=1989) | Compared to continuous supply |  | Compared to intermittent supply meeting SDG |  | Intermittent supply meeting SDG (N=229) | Compared to continuous supply |  |
|  | Mean (SD) | aCIR (95% CI) | p-value | aCIR (95% CI) | p-value | Mean (SD) | aCIR (95% CI) | p-value |
| Typhoid fever | 0.09 (0.42) | **1.93 (1.15, 3.22)** | **0.01** | **3.66 (1.37, 9.79)** | **0.01** | 0.03 (0.19) | 0.54 (0.20, 1.45) | 0.22 |
| Confirmed typhoid fever | 0.07 (0.33) | **1.85 (1.12, 3.05)** | **0.01** | **3.60 (1.57, 8.30)** | **<0.005** | 0.02 (0.17) | 0.54 (0.23, 1.24) | 0.15 |

aPR: Adjusted prevalence ratio; aCIR: Adjusted cumulative incidence ratio; CI: Confidence interval; SD: Standard deviation

### Intermittent supply frequency and duration vs. continuous supply

Among IWS households in the upper quintiles of supply frequency and duration (those receiving water at shorter intervals and/or for more hours per supply cycle), waterborne risks were broadly indistinguishable from CWS households (Figs 2 and 3, Table S2). In contrast, IWS households in the lower quintiles of supply interval and duration (those receiving water less often and/or for fewer hours per supply cycle) had significantly higher prevalence of child diarrhea and incidence of typhoid fever than CWS households (Figs 2 and 3, Table S2). Within IWS households, the prevalence of diarrhea among children <5 years and the incidence of typhoid fever both increased progressively with increasing number of days between supply cycles and decreasing hours of supply (Figs 2 and 3, Table S3).

**Fig 2.**
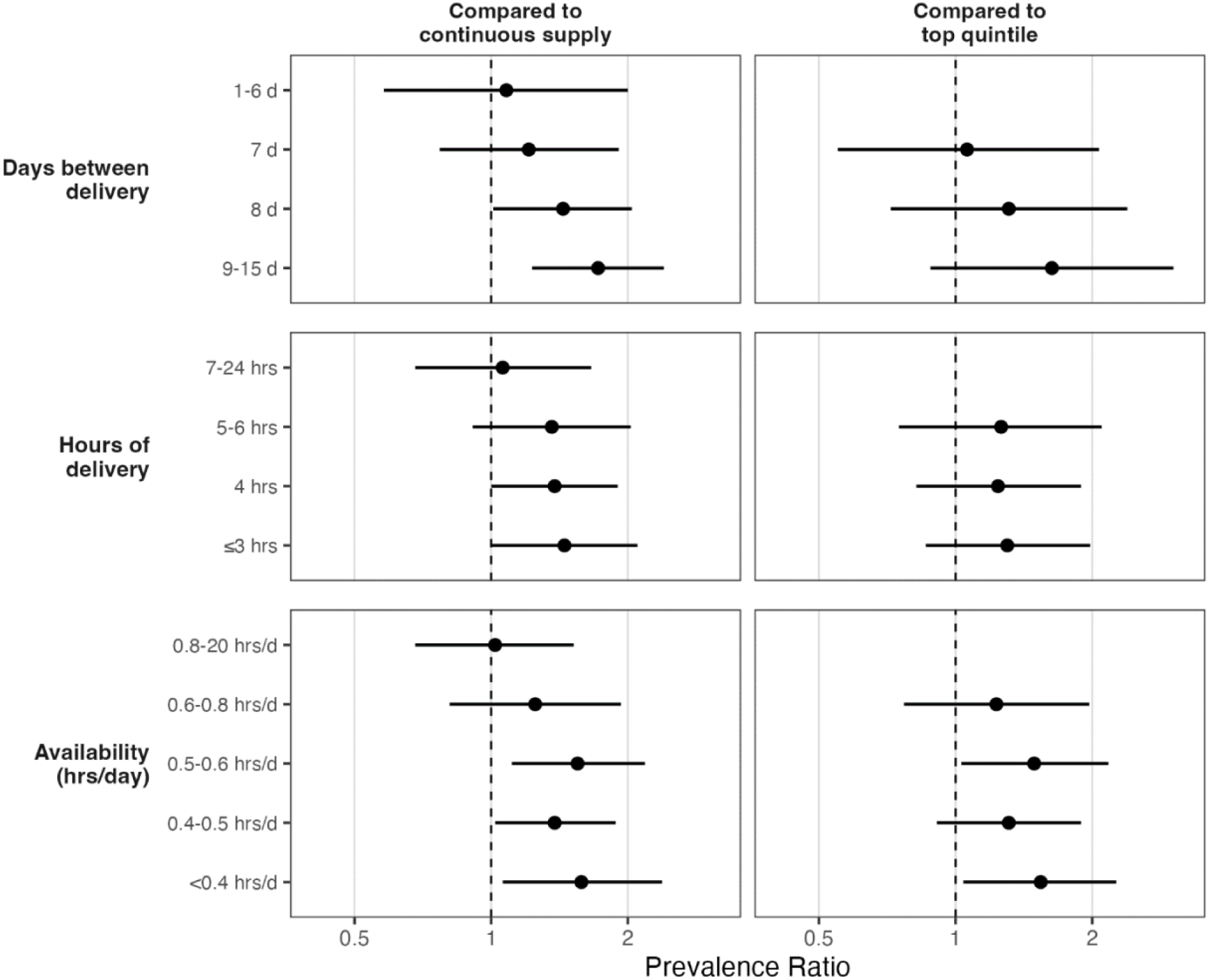
Prevalence ratios for diarrhea in children <5 years across quintiles of water supply frequency (days between delivery), duration (hours of delivery) and availability (average hours of delivery per day). Ratios on the left compare all quintiles of intermittent water supply to continuous water supply; ratios on the right compare the lower quintiles of intermittent water supply (less frequent/shorter delivery) to the top quintile of intermittent water supply. Circles denote point estimates; horizontal lines denote 95% confidence intervals.

**Fig 3.**
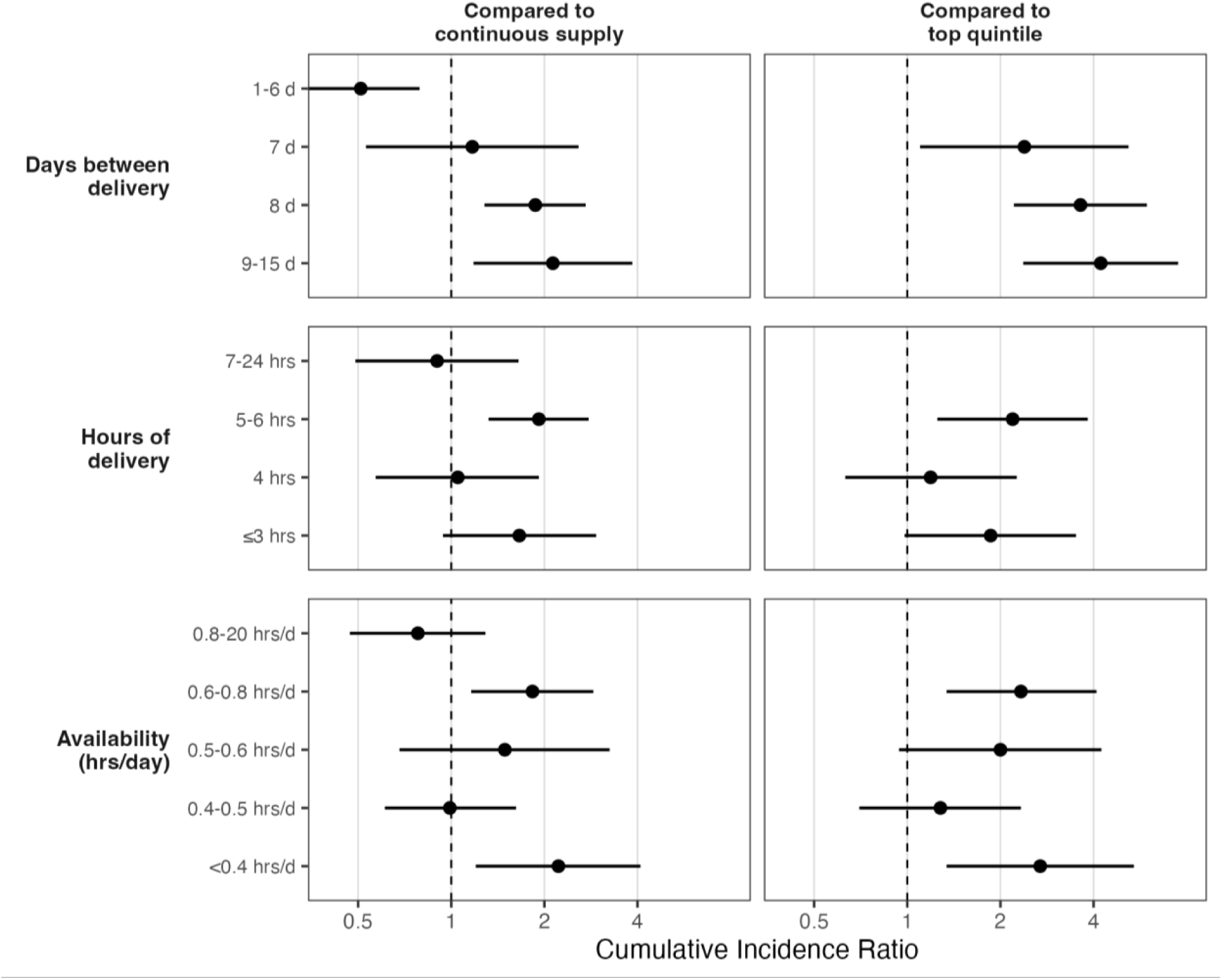
Cumulative incidence ratio for typhoid fever among household members across quintiles of water supply frequency (days between delivery), duration (hours of delivery) and availability (average hours of delivery per day). Ratios on the left compare all quintiles of intermittent water supply to continuous water supply; ratios on the right compare the lower quintiles of intermittent water supply (less frequent/shorter delivery) to the top quintile of intermittent water supply. Circles denote point estimates; horizontal lines denote 95% confidence intervals.

#### Supply frequency

Compared to CWS, IWS households in the top quintile of supply interval (receiving water once every 1-6 days) had similar prevalence of child diarrhea (PR=1.08, 0.58-2.00), 49% lower incidence of ≥1 typhoid fever case in the household (CIR=0.51, 0.33-0.79) and 49% fewer typhoid fever cases (CIR=0.51, 0.30-0.87) (Figs 2 and 3, Table S2). Compared to CWS, IWS households in the bottom quintile of supply interval (receiving water once every 9-15 days) had 72% higher prevalence of child diarrhea (PR=1.72, 1.23-2.40), 2 times higher incidence of ≥1 typhoid fever case in the household (CIR=2.13, 1.18-3.85) and twice as many typhoid fever cases (CIR=1.96, 1.11-3.47) (Figs 2 and 3, Table S2). Compared to IWS households in the top quintile of supply interval, those in the bottom quintile also appeared to have 63% higher prevalence of child diarrhea (PR=1.63, 0.88-3.02), and had 4 times higher incidence of ≥1 typhoid fever case in the household (CIR=4.22, 2.37-7.51) and 4 times as many typhoid fever cases (CIR=3.77, 1.88-7.54) than (Figs 2 and 3, Table S3).

#### Supply duration

Among IWS households in the top quintile of supply duration (receiving water for 7-24 hours per supply cycle), the prevalence of child diarrhea and the incidence of typhoid fever was indistinguishable from CWS households (p>0.05) (Figs 2 and 3, Table S2). In contrast, IWS households in the bottom quintile of supply duration (receiving water for ≤3 hours per supply cycle) had 45% higher prevalence of child diarrhea (PR=1.45, 1.00-2.10), 66% higher incidence of ≥1 typhoid fever case in the household (CIR=1.66, 0.94-2.94), and twice as many typhoid fever cases (CIR=1.96, 1.11-3.47) than those with CWS (Figs 2 and 3, Table S2). Compared to households in the top quintile of supply duration, those in the bottom quintile appeared to have 2 times higher incidence of ≥1 typhoid fever case in the household (CIR=1.86, 0.98-3.51) and had twice as many typhoid fever cases (CIR=2.33, 1.35-4.02) than the top quintile (Figs 2 and 3, Table S3).

#### Average supply availability

Among IWS households in the top quintile of supply availability (receiving water for 0.8-20 hours/day on average), the prevalence of child diarrhea and the incidence of typhoid fever was indistinguishable from CWS households (p>0.05) (Figs 2 and 3, Table S2). In contrast, IWS households in the bottom quintile of supply availability (receiving water for <0.4 hours/day on average) had 58% higher prevalence of child diarrhea (PR=1.58, 1.06-2.38), over 2 times higher incidence of ≥1 typhoid fever case in the household (CIR=2.22, 1.20-4.09), and 2.5 times as many typhoid fever cases (CIR=2.55, 1.42-4.61) than households with CWS (Figs 2 and 3, Table S2). Compared to households in the top quintile of supply availability, those in the bottom quintile had 54% higher prevalence of child diarrhea (PR=1.54, 1.04-2.26), almost 3 times higher incidence of ≥1 typhoid fever case in the household (CIR=2.69, 1.34-5.40) and 3 times as many typhoid fever cases (CIR=3.25, 1.70-6.20) (Figs 2 and 3, Table S3).

Findings were similar for the prevalence of diarrhea or blood in stool and for the incidence of laboratory-confirmed typhoid fever (Tables S2-S3). The 7-day prevalence of ear infection among children <5 years in IWS households in the bottom quintile of supply duration was 42% higher than those in CWS households and 69% higher those in the top quintile (p-values<0.05, Table S4). Children in IWS households in the bottom quintile of average supply availability also had 46% higher prevalence of ear infection that those in CWS households (p-value<0.05, Table S4). There was no association between ear infections and any other quintile of supply interval, duration, or availability.

## Discussion

Approximately a decade after the initiation of CWS in Hubli-Dharwad, we found sustained health benefits for households with continuous supply: children <5 years with IWS had 36% higher diarrhea than those with CWS in our current study. This aligns with our previous evaluation conducted in the same population approximately three years after the initiation of CWS, which found that lower-income IWS households had 59% higher prevalence of severe enteric infections (i.e. blood in stool) in children <5 years than lower-income CWS households [23]. The current study enrolled from among households with no weightbearing roof to rule out ongoing water supply from overhead storage tanks during intermittencies. Therefore, we expect that we enrolled lower-income households, and our findings align with our previous evaluation.

In both evaluations, IWS was associated with increased risk of typhoid fever. Outbreaks of typhoid fever have been associated with deficiencies in piped water supplies, such as lack of chlorine residual and leaky water pipes [8,31]. In our previous evaluation, we detected free chlorine residual at recommended levels (>0.2 mg/L) in 40% of water samples from IWS households vs. 68% of samples from CWS households [25]. Our findings corroborate that pathogen intrusion into pipes in intermittent systems can drive *Salmonella typhi* transmission. Given increasing trends of drug resistance in *Salmonella typhi,* including multidrug-resistant (MDR) and extensively drug-resistant (XDR) strains [32], intermittent water supplies could be a risk factor for antimicrobial-resistant *Salmonella typhi* infections.

This study also adds nuance to our understanding of health risks associated with different manifestations of IWS. When we evaluated the extent of intermittency among IWS households, IWS households that met the SDG criterion of having water supply for at least 4 days/week or 12 hours/day had health outcomes similar to CWS households. Similarly, among IWS households in the top quintile of water supply frequency and duration (those receiving water every ≤6 days and for ≥7 hours per supply cycle), waterborne risks were indistinguishable from CWS households. In contrast, IWS households that fell below the SDG criterion and those in the lower quintiles of water supply frequency and duration (receiving water every ≥9 days and for ≤3 hours per supply cycle) experienced significantly more child diarrhea and typhoid fever than CWS households. Within IWS households, the prevalence of child diarrhea and incidence of typhoid fever both increased with increasing number of days between supply cycles and decreasing hours of supply. This is consistent with our recent re-analysis of the 2010-2011 data from Hubli-Dharwad, demonstrating that health risks from water supply unpredictability among IWS households were significantly higher for households with infrequent supply, aligning with the bottom-quintile findings in the current analysis: children are at greatest risk of waterborne illness when households receive water infrequently and cannot predict when they will receive it. Taken together, our findings indicate that, while delivering a continuous piped water supply is the overarching water security goal, efforts to increase service levels of continuity, such as shorter supply intervals and longer supply duration [33], can help reduce waterborne disease risks among populations served by intermittent systems. The JMP metric for meeting the UN SDGs aims for “water available when needed” – at least 12 hours per day or 4 days per week [21]; our findings indicate that meeting this criterion is associated with improved health and also highlight that less strict thresholds still provided protection. Notably, as of 2026, approximately 20 years after the initiation of the upgrade from IWS to CWS, wards with continuous supply in Hubli-Dharwad are still limited. Therefore, while continuous supply has been sustained in the wards where it was initiated, it has not expanded to the entire city, questioning the feasibility of scale and corroborating the importance of improving supply frequency and duration in intermittent supplies while working towards achieving continuous supplies.

Our study had limitations. We expect that matching in the design phase and controlling for potential confounders in the analysis phase reduced confounding, but residual confounding may remain. Our outcomes were self-reported and could be susceptible to information bias. However, our negative control outcome, ear infections among children, generally did not show associations with the exposure variables in our analysis, except for increased prevalence of ear infection among the bottom water supply duration and availability quintiles; these increases were not associated with any dose-response relationships and were only observed in the bottom quintiles of these two variables, while the diarrhea and typhoid fever outcomes showed dose-response patterns with increasing severity of intermittency. Therefore, while confounding or information bias may have affected some of our findings, the lack of associations with the negative control outcome for most exposure variables indicates no overall bias across the study. Additionally, findings were similar when we limited the analysis to cases of typhoid fever that were reported to be lab-confirmed vs. reported to be diagnosed based on symptoms, indicating no information bias. Finally, our study period overlapped with the dry season in Hubli-Dharwad; therefore, our findings may not generalize to wet periods, such as the monsoon season.

In our study in an urban Indian setting with piped water supply, we found lower risk of waterborne disease associated with incremental improvements in water supply frequency and duration, along the continuum from infrequent/short intermittent delivery to frequent/long intermittent delivery to continuous delivery. Other recent findings showed that predictable intermittent supplies achieved similar health benefits as continuous supplies; this current study shows that providing supply more often or longer can also close the gap and achieve health benefits that are similar to continuous supply. These findings support global efforts to implement water supply improvements that focus on improving the level of service in terms of supply continuity; however, importantly, our results highlight that increasing how often and for how many hours supplies are delivered in intermittent systems in the interim can deliver health benefits and the UN criterion of having “water available when needed” improves health.

## Data Availability

All data produced will be available online at Open Science Framework upon publication.

## Supplemental Materials

Characteristics of enrolled households; associations between water supply characteristics and waterborne infections (compared to continuous supply); associations between water supply characteristics and waterborne infections (compared to top quintile of intermittent supply frequency/duration); associations between water supply characteristics and negative control outcome.

## Acknowledgements

This study was funded by the University of Massachusetts, Amherst Civil and Environmental Engineering Department.

## Notes

### Competing Interest Statement

The authors have declared no competing interest.

### Author Declarations

The Institutional Review Board of University of Massachusetts, Amherst gave ethical approval for this work (2018-5063), with an Institutional Reliance Agreement (21033) by North Carolina State University.

