## Supplemental Materials for "Water Supply Continuity, Frequency, and Health Gains: Ten-year Evidence from Hubli-Dharwad, India"

<u>**Table of Contents**</u>

**Table S1.** Characteristics of enrolled households by study arm

**Table S2.** Associations between water supply characteristics and waterborne infections, compared to continuous supply

**Table S3.** Associations between water supply characteristics and waterborne infections, compared to top quintile of supply frequency/duration

**Table S4.** Associations between water supply characteristics and the prevalence of ear infection (negative control outcome)

**Table S1.** Characteristics of enrolled households by study arm

| | All households N=4438 | Intermittent supply N=2218 | Continuous supply N=2220 |
|---|---|---|---|
| Socio-demographics | | | |
| Number of household members, mean (SD) | 6.4 (3.9) | 6.3 (3.9) | 6.5 (3.9) |
| Age of children <5 yrs, mean (SD) | 2.1 (1.4) | 2.2 (1.4) | 2.1 (1.4) |
| Age of mother of children <5 yrs, mean (SD) | 26.4 (3.8) | 26.0 (3.6) | 26.7 (4.0) |
| Education of mother of children <5 yrs, % (n) | | | |
| No schooling | 12.2 (540) | 11.8 (260) | 12.6 (280) |
| Primary | 34.3 (1518) | 38.4 (849) | 30.1 (669) |
| Secondary | 42.0 (1862) | 40.3 (892) | 43.7 (970) |
| University or above | 11.5 (511) | 9.5 (210) | 13.6 (301_ |
| Religion, % (n) | | | |
| Hindu | 63.6 (2822) | 65.0 (1441) | 62.2 (1381) |
| Muslim | 35.0 (1552) | 33.7 (747) | 36.3 (805) |
| Other | 1.4 (63) | 1.3 (29) | 1.5 (34) |
| Scheduled caste or tribe, % (n) | 22.5 (998) | 21.6 (477) | 23.6 (521) |
| Household holds ration card, % (n) [a] | 88.5 (3926) | 88.5 (1954) | 89.1 (1972) |
| Household owns their home, % (n) | 73.5 (3260) | 73.4 (1627) | 73.6 (1633) |
| Chawl house, % (n) [b] | 9.0 (401) | 8.7 (193) | 9.4 (208) |
| Number of rooms in home | 1.8 (0.9) | 1.8 (1.0) | 1.8 (0.8) |
| Floor material, % (n) | | | |
| Stone/cement/tile | 97.7 (4336) | 97.2 (2155) | 98.2 (2181) |
| Mud/wood | 2.3 (102) | 2.8 (63) | 1.8 (39) |
| Wall material, % (n) | | | |
| Brick/concrete/stone | 72.6 (3221) | 72.2 (1602) | 72.9 (1619) |
| Mud/wood/metal | 27.4 (1217) | 27.8 (616) | 27.1 (601) |
| Roof material, % (n) | | | |
| Tiled | 49.1 (2180) | 45.4 (1006) | 52.3 (1174) |
| Mud/wood/metal | 50.9 (2258) | 54.6 (1212) | 47.1 (1046) |
| Household owns, % (n) | | | |
| Fridge | 13.4 (595) | 12.7 (282) | 14.1 (313) |
| TV | 93.9 (4168) | 92.4 (2050) | 95.4 (2118) |
| Mobile phone | 68.5 (3041) | 69.8 (1548) | 67.3 (1493) |
| Smart phone | 76.3 (3387) | 72.6 (1610) | 80.1 (1777) |
| Two-wheeler | 53.3 (2364) | 54.0 (1197) | 52.6 (1167) |
| Wealth index, % (n) | | | |
| Top quartile | 25.0 (1100) | 27. 6 (611) | 22.5 (499) |
| 2nd quartile | 25.0 (1109) | 24.0 (532) | 26.0 (577) |
| 3rd quartile | 25.9 (1149) | 24.2 (536) | 27.6 (613) |
| Bottom quartile | 24.1 (1070) | 24.3 (539) | 23.9 (531) |
| Sanitation and hygiene conditions | | | |
| Toilet access, % (n) | | | |
| Non-shared private | 82.7 (3670) | 80.0 (1771) | 85.5 (1899) |
| Shared private | 10.9 (484) | 12.7 (281) | 9.1 (203) |
| Public | 2.3 (100) | 2.4 (54) | 2.1 (46) |
| None | 4.2 (184) | 5.1 (112) | 3.2 (72) |
| Open drain adjacent to home, % (n) [c] | 57.8 (2565) | 62.0 (1373) | 53.7 (1192) |
| Soap/detergent present in toilet area, % (n) | 30.2 (1340) | 27.1 (577) | 35.6 (763) |
| Soap/detergent present in kitchen area, % (n) | 10.7 (474) | 7.6 (165) | 14.3 (309) |

SD: Standard deviation.

[a] Ration card is given to low-income households to provide food subsidies.

[b] Chawl house refers to low-cost multi-family housing unit where each family typically occupies one room.

[c] Adjacent defined as on/at the edge of their property or on their street.

**Table S2.** Associations between water supply characteristics and waterborne infections, compared to continuous supply

| Prevalence in children <5 years: | | | Diarrhea | | | Diarrhea or blood | | |
|---|---|---|---|---|---|---|---|---|
| Supply quintile | Range | N | % (n) | aPR (95% CI) | p-value | % (n) | aPR (95% CI) | p-value |
| Days between water delivery | | | | | | | | |
| Continuous | 0 days | 3172 | 5.5 (174) | ref | -- | 6.0 (189) | -- | -- |
| 1 | 1-6 days | 641 | 6.2 (40) | 1.08 (0.58, 2.00) | 0.81 | 7.0 (45) | 1.13 (0.62, 2.05) | 0.69 |
| 2 | 7 days | 690 | 7.1 (49) | 1.21 (0.77, 1.91) | 0.41 | 7.8 (54) | 1.27 (0.85, 1.89) | 0.25 |
| 3 | 8 days | 1480 | 7.6 (112) | **1.44 (1.01, 2.04)** | **0.04** | 7.8 (15) | **1.39 (1.00, 1.92)** | **0.05** |
| 5 | 9-15 days | 368 | 9.0 (33) | **1.72 (1.23, 2.40)** | **0.002** | 9.2 (34) | **1.64 (1.19, 2.26)** | **0.002** |
| Hours of water delivery | | | | | | | | |
| Continuous | 24 hrs | 3172 | 5.5 (174) | ref | -- | 6.0 (189) | -- | -- |
| 1 | 7-24 hrs | 463 | 5.6 (26) | 1.06 (0.68, 1.66) | 0.80 | 6.1 (463) | 1.08 (0.72, 1.63) | 0.71 |
| 2 | 5-6 hrs | 677 | 7.8 (53) | 1.36 (0.91, 2.03) | 0.13 | 8.4 (57) | 1.37 (0.94, 1.98) | 0.10 |
| 4 | 4 hrs | 874 | 7.3 (64) | **1.38 (1.00, 1.90)** | **0.05** | 7.8 (68) | **1.36 (1.01, 1.83)** | **0.04** |
| 5 | ≤3 hrs | 1164 | 7.8 (91) | **1.45 (1.00, 2.10)** | **0.05** | 8.2 (95) | 1.41 (0.98, 2.02) | 0.06 |
| Water availability (hours/day) | | | | | | | | |
| Continuous | 24 hrs/day | 3172 | 5.5 (174) | ref | -- | 6.0 (189) | -- | -- |
| 1 | 0.8-20 hrs/day | 623 | 5.5 (34) | 1.02 (0.68, 1.52) | 0.94 | 6.3 (39) | 1.08 (0.73, 1.60) | 0.71 |
| 2 | 0.6-0.8 hrs/day | 602 | 7.3 (44) | 1.25 (0.81, 1.93) | 0.31 | 7.8 (47) | 1.25 (0.84, 1.85) | 0.27 |
| 3 | 0.5-0.6 hrs/day | 377 | 8.5 (32) | **1.55 (1.11, 2.18)** | **0.01** | 9.6 (36) | **1.64 (1.22, 2.22)** | **<0.005** |
| 4 | 0.4-0.5 hrs/day | 900 | 7.4 (67) | **1.38 (1.02, 1.88)** | **0.04** | 7.6 (68) | 1.32 (0.98, 1.78) | 0.07 |
| 5 | <0.4 hrs/day | 676 | 8.4 (57) | **1.58 (1.06, 2.38)** | **0.03** | 8.6 (58) | **1.51 (1.03, 2.21)** | **0.04** |
| Households with at least one case: | | | Typhoid fever | | | Confirmed typhoid fever | | |
| Supply quintile | Range | N | % (n) | aCIR (95% CI) | p-value | % (n) | aCIR (95% CI) | p-value |
| Days between water delivery | | | | | | | | |
| Continuous | 0 days | 2220 | 4.3 (95) | ref | -- | 3.3 (73) | -- | -- |
| 1 | 1-6 days | 456 | 2.6 (12) | **0.51 (0.33, 0.79)** | **<0.005** | 2.0 (9) | **0.49 (0.32, 0.77)** | **<0.005** |
| 2 | 7 days | 491 | 6.3 (31) | 1.17 (0.53, 2.58) | 0.70 | 4.9 (24) | 1.11 (0.54, 2.28) | 0.77 |
| 3 | 8 days | 1021 | 7.9 (81) | **1.87 (1.28, 2.72)** | **<0.005** | 6.3 (64) | **1.93 (1.25, 2.97)** | **<0.005** |
| 5 | 9-15 days | 250 | 7.6 (19) | **2.13 (1.18, 3.85)** | **0.01** | 6.4 (16) | **2.33 (1.17, 4.63)** | **0.02** |
| Hours of water delivery | | | | | | | | |
| Continuous | 24 hrs | 2220 | 4.3 (95) | ref | -- | 3.3 (73) | -- | -- |
| 1 | 7-24 hrs | 340 | 3.8 (13) | 0.90 (0.49, 1.65) | 0.72 | 3.2 (11) | 0.94 (0.46, 1.91) | 0.87 |
| 2 | 5-6 hrs | 440 | 8.9 (39) | **1.92 (1.32, 2.78)** | **<0.005** | 7.3 (32) | **1.96 (1.22, 3.16)** | **0.01** |
| 4 | 4 hrs | 606 | 4.8 (29) | 1.05 (0.57, 1.92) | 0.88 | 3.1 (19) | 0.89 (0.50, 1.57) | 0.68 |
| 5 | ≤3 hrs | 831 | 7.5 (62) | 1.66 (0.94, 2.94) | 0.08 | 6.1 (51) | 1.76 (0.92, 3.35) | 0.09 |
| Water availability (hours/day) | | | | | | | | |
| Continuous | 24 hrs/day | 2220 | 4.3 (95) | ref | -- | 3.3 (73) | -- | -- |
| 1 | 0.8-20 hrs/day | 449 | 3.6 (16) | 0.78 (0.47, 1.29) | 0.33 | 3.3 (15) | 0.91 (0.53, 1.56) | 0.73 |
| 2 | 0.6-0.8 hrs/day | 398 | 8.9 (35) | **1.83 (1.16, 2.88)** | **0.01** | 7.0 (28) | **1.87 (1.07, 3.26)** | **0.03** |
| 3 | 0.5-0.6 hrs/day | 254 | 7.5 (19) | 1.49 (0.68, 3.25) | 0.232 | 5.1 (13) | 1.29 (0.61, 2.71) | 0.50 |
| 4 | 0.4-0.5 hrs/day | 628 | 4.6 (29) | 0.99 (0.61, 1.62) | 0.98 | 3.3 (21) | 0.89 (0.49, 1.61) | 0.69 |
| 5 | <0.4 hrs/day | 488 | 9.0 (44) | **2.22 (1.20, 4.09)** | **0.01** | 7.4 (36) | **2.38 (1.17, 4.84)** | **0.02** |
| Number of cases per household: | | | Typhoid fever | | | Confirmed typhoid fever | | |
| Supply quintile | Range | N | Mean (SD) | aCIR (95% CI) | p-value | Mean (SD) | aCIR (95% CI) | p-value |
| Days between water delivery | | | | | | | | |
| Continuous | 0 days | 2220 | 0.05 (0.23) | ref | -- | 0.04 (0.21) | ref | -- |
| 1 | 1-6 days | 456 | 0.03 (0.18) | **0.51 (0.30, 0.87)** | **0.02** | 0.02 (0.16) | **0.49 (0.28, 0.85)** | **0.01** |
| 2 | 7 days | 491 | 0.11 (.60) | 2.02 (0.61, 6.74) | 0.25 | 0.08 (0.40) | 1.58 (0.60, 4.17) | 0.35 |
| 3 | 8 days | 1021 | 0.10 (0.38) | **2.23 (1.49, 3.33)** | **<0.005** | 0.08 (0.35) | **2.36 (1.48, 3.77)** | **<0.005** |
| 5 | 9-15 days | 250 | 0.08 (0.27) | **1.96 (1.05, 3.66)** | **0.03** | 0.06 (0.25) | **2.12 (1.04, 4.33)** | **0.04** |
| Hours of water delivery | | | | | | | | |
| Continuous | 24 hrs | 2220 | 0.05 (0.23) | ref | -- | 0.04 (0.21) | ref | -- |
| 1 | 7-24 hrs | 340 | 0.04 (0.21) | 0.87 (0.48, 1.58) | 0.65 | 0.04 (0.20) | 0.92 (0.48, 1.76) | 0.80 |
| 2 | 5-6 hrs | 440 | 0.11 (0.41) | **2.27 (1.53, 3.37)** | **<0.005** | 0.10 (0.40) | **2.41 (1.47, 3.97)** | **<0.005** |
| 4 | 4 hrs | 606 | 0.08 (0.49) | 1.68 (0.59, 4.79) | 0.33 | 0.04 (0.28) | 1.15 (0.52, 2.51) | 0.73 |
| 5 | ≤3 hrs | 831 | 0.09 (0.37) | **1.96 (1.11, 3.47)** | **0.02** | 0.08 (0.35) | **2.05 (1.06, 3.97)** | **0.03** |
| Water availability (hours/day) | | | | | | | | |
| Continuous | 24 hrs/day | 2220 | 0.05 (0.23) | ref | -- | 0.04 (0.21) | ref | -- |
| 1 | 0.8-20 hrs/day | 449 | 0.04 (0.20) | 0.76 (0.45, 1.28) | 0.30 | 0.04 (0.20) | 0.87 (0.52, 1.47) | 0.61 |
| 2 | 0.6-0.8 hrs/day | 398 | 0.11 (0.43) | **2.22 (1.38, 3.56)** | **<0.005** | 0.10 (0.41) | **2.34 (1.32, 4.16)** | **<0.005** |
| 3 | 0.5-0.6 hrs/day | 254 | 0.14 (0.72) | 2.70 (0.78, 9.36) | 0.12 | 0.08 (0.40) | 1.80 (0.73, 4.43) | 0.20 |
| 4 | 0.4-0.5 hrs/day | 628 | 0.06 (0.32) | 1.18 (0.69, 2.04) | 0.55 | 0.04 (0.27) | 1.00 (0.55, 1.83) | 0.99 |
| 5 | <0.4 hrs/day | 488 | 0.11 (0.38) | **2.55 (1.42, 4.61)** | **<0.005** | 0.09 (0.35) | **2.79 (1.41, 5.54)** | **<0.005** |

aPR: Adjusted prevalence ratio; aCIR: Adjusted cumulative incidence ratio; CI: Confidence interval; SD: Standard deviation

**Table S3.** Associations between water supply characteristics and waterborne infections, compared to top quintile of supply frequency/duration

| Prevalence in children <5 years: | | | Diarrhea | | | Diarrhea or blood | | |
|---|---|---|---|---|---|---|---|---|
| Supply quintile | Range | N | % (n) | aPR (95% CI) | p-value | % (n) | aPR (95% CI) | p-value |
| Days between water delivery | | | | | | | | |
| 1 | 1-6 days | 641 | 6.2 (40) | ref | -- | 7.0 (45) | ref | -- |
| 2 | 7 days | 690 | 7.1 (49) | 1.06 (0.55, 2.07) | 0.85 | 7.8 (54) | 1.07 (0.59, 1.96) | 0.82 |
| 3 | 8 days | 1480 | 7.6 (112) | 1.31 (0.72, 2.39) | 0.38 | 7.8 (15) | 1.21 (0.68, 2.15) | 0.61 |
| 5 | 9-15 days | 368 | 9.0 (33) | 1.63 (0.88, 3.02) | 0.12 | 9.2 (34) | 1.47 (0.81, 2.68) | 0.21 |
| Hours of water delivery | | | | | | | | |
| 1 | 7-24 hrs | 463 | 5.6 (26) | ref | -- | 6.1 (463) | ref | -- |
| 2 | 5-6 hrs | 677 | 7.8 (53) | 1.26 (0.75, 2.10) | 0.39 | 8.4 (57) | 1.24 (0.79,1.95) | 0.35 |
| 4 | 4 hrs | 874 | 7.3 (64) | 1.24 (0.82, 1.89) | 0.31 | 7.8 (68) | 1.21 (0.81, 1.81) | 0.36 |
| 5 | ≤3 hrs | 1164 | 7.8 (91) | 1.30 (0.86, 1.98) | 0.22 | 8.2 (95) | 1.26 (0.86, 1.84) | 0.24 |
| Water availability (hours/day) | | | | | | | | |
| 1 | 0.8-20 hrs/day | 623 | 5.5 (34) | ref | -- | 6.3 (39) | ref | -- |
| 2 | 0.6-0.8 hrs/day | 602 | 7.3 (44) | 1.23 (0.77, 1.97) | 0.38 | 7.8 (47) | 1.17 (0.78, 1.73) | 0.45 |
| 3 | 0.5-0.6 hrs/day | 377 | 8.5 (32) | **1.49 (1.03, 2.17)** | **0.03** | 9.6 (36) | **1.49 (1.00, 2.22)** | **0.05** |
| 4 | 0.4-0.5 hrs/day | 900 | 7.4 (67) | 1.31 (0.91, 1.89) | 0.14 | 7.6 (68) | 1.19 (0.84, 1.67) | 0.33 |
| 5 | <0.4 hrs/day | 676 | 8.4 (57) | **1.54 (1.04, 2.26)** | **0.03** | 8.6 (58) | 1.39 (0.96, 2.01) | 0.08 |
| Households with at least one case: | | | Typhoid fever | | | Confirmed typhoid fever | | |
| Supply quintile | Range | N | % (n) | aCIR (95% CI) | p-value | % (n) | aCIR (95% CI) | p-value |
| Days between water delivery | | | | | | | | |
| 1 | 1-6 days | 456 | 2.6 (12) | ref | -- | 2.0 (9) | ref | -- |
| 2 | 7 days | 491 | 6.3 (31) | **2.39 (1.10, 5.19)** | **0.03** | 4.9 (24) | **2.35 (1.20, 4.61)** | **0.01** |
| 3 | 8 days | 1021 | 7.9 (81) | **3.63 (2.21, 5.95)** | **<0.005** | 6.3 (64) | **3.91 (2.27, 6.72)** | **<0.005** |
| 5 | 9-15 days | 250 | 7.6 (19) | **4.22 (2.37, 7.51)** | **<0.005** | 6.4 (16) | **4.71 (2.47, 8.97)** | **<0.005** |
| Hours of water delivery | | | | | | | | |
| 1 | 7-24 hrs | 340 | 3.8 (13) | ref | -- | 3.2 (11) | ref | -- |
| 2 | 5-6 hrs | 440 | 8.9 (39) | **2.19 (1.25, 3.83)** | **0.01** | 7.3 (32) | **2.13 (1.09, 4.58)** | **0.03** |
| 4 | 4 hrs | 606 | 4.8 (29) | 1.19 (0.63, 2.26) | 0.58 | 3.1 (19) | 0.96 (0.47, 1.97) | 0.92 |
| 5 | ≤3 hrs | 831 | 7.5 (62) | 1.86 (0.98, 3.51) | 0.06 | 6.1 (51) | 1.84 (0.89, 3.82) | 0.10 |
| Water availability (hours/day) | | | | | | | | |
| 1 | 0.8-20 hrs/day | 449 | 3.6 (16) | ref | -- | 3.3 (15) | ref | -- |
| 2 | 0.6-0.8 hrs/day | 398 | 8.9 (35) | **2.33 (1.34, 4.09)** | **<0.005** | 7.0 (28) | **2.08 (1.14, 3.78)** | **0.02** |
| 3 | 0.5-0.6 hrs/day | 254 | 7.5 (19) | 2.00 (0.94, 4.24) | 0.07 | 5.1 (13) | 1.48 (0.69, 3.20) | 0.32 |
| 4 | 0.4-0.5 hrs/day | 628 | 4.6 (29) | 1.28 (0.70, 2.33) | 0.42 | 3.3 (21) | 0.98 (0.47, 2.03) | 0.95 |
| 5 | <0.4 hrs/day | 488 | 9.0 (44) | **2.69 (1.34, 5.40)** | **0.01** | 7.4 (36) | **2.47 (1.11, 5.45)** | **0.03** |
| Number of cases per household: | | | Typhoid fever | | | Confirmed typhoid fever | | |
| Supply quintile | Range | N | Mean (SD) | aCIR (95% CI) | p-value | Mean (SD) | aCIR (95% CI) | p-value |
| Days between water delivery | | | | | | | | |
| 1 | 1-6 days | 456 | 0.03 (0.18) | ref | -- | 0.02 (0.16) | ref | -- |
| 2 | 7 days | 491 | 0.11 (.60) | **4.12 (1.15, 14.7)** | **0.03** | 0.08 (0.40) | **3.41 (1.20, 9.75)** | **0.02** |
| 3 | 8 days | 1021 | 0.10 (0.38) | **4.35 (2.39, 7.90)** | **<0.005** | 0.08 (0.35) | **5.02 (2.58, 9.77)** | **<0.005** |
| 5 | 9-15 days | 250 | 0.08 (0.27) | **3.77 (1.88, 7.54)** | **<0.005** | 0.06 (0.25) | **4.31 (2.03, 9.15)** | **<0.005** |
| Hours of water delivery | | | | | | | | |
| 1 | 7-24 hrs | 340 | 0.04 (0.21) | ref | -- | 0.04 (0.20) | ref | -- |
| 2 | 5-6 hrs | 440 | 0.11 (0.41) | **2.72 (1.59, 4.66)** | **<0.005** | 0.10 (0.40) | **2.76 (1.50, 5.08)** | **<0.005** |
| 4 | 4 hrs | 606 | 0.08 (0.49) | 2.02 (0.72, 5.69) | 0.18 | 0.04 (0.28) | **1.31 (0.59, 2.94)** | **0.51** |
| 5 | ≤3 hrs | 831 | 0.09 (0.37) | **2.33 (1.35, 4.02)** | **<0.005** | 0.08 (0.35) | **2.29 (1.26, 4.16)** | **0.01** |
| Water availability (hours/day) | | | | | | | | |
| 1 | 0.8-20 hrs/day | 449 | 0.04 (0.20) | ref | -- | 0.04 (0.20) | ref | -- |
| 2 | 0.6-0.8 hrs/day | 398 | 0.11 (0.43) | **2.92 (1.69, 5.04)** | **<0.005** | 0.10 (0.41) | **2.73 (1.53, 4.87)** | **<0.005** |
| 3 | 0.5-0.6 hrs/day | 254 | 0.14 (0.72) | **3.74 (1.17, 11.9)** | **0.03** | 0.08 (0.40) | 2.16 (0.94, 4.94) | 0.07 |
| 4 | 0.4-0.5 hrs/day | 628 | 0.06 (0.32) | 1.58 (0.88, 2.86) | 0.13 | 0.04 (0.27) | 1.18 (0.63, 2.19) | 0.61 |
| 5 | <0.4 hrs/day | 488 | 0.11 (0.38) | **3.25 (1.70, 6.20)** | **<0.005** | 0.09 (0.35) | **3.13 (1.55, 6.30)** | **<0.005** |

aPR: Adjusted prevalence ratio; aCIR: Adjusted cumulative incidence ratio; CI: Confidence interval; SD: Standard deviation

**Table S4.** Associations between water supply characteristics and the prevalence of ear infection (negative control outcome)

| | | | Ear infection prevalence | Compared to continuous | | Compared to top quintile | |
|---|---|---|---|---|---|---|---|
| Supply quintile | Range | N | % (n) | aPR (95% CI) | p-value | aPR (95% CI) | p-value |
| Days between water delivery | | | | | | | |
| Continuous | 0 days | 3172 | 6.7 (211) | ref | -- | | |
| 1 | 1-6 days | 641 | 6.6 (42) | 0.92 (0.65, 1.31) | 0.65 | ref | -- |
| 2 | 7 days | 690 | 8.3 (57) | 1.25 (0.77, 2.03) | 0.38 | 1.38 (0.84, 2.25) | 0.20 |
| 3 | 8 days | 1480 | 7.0 (103) | 1.08 (0.74, 1.56) | 0.70 | 1.14 (0.79, 1.64) | 0.47 |
| 5 | 9-15 days | 368 | 6.3 (23) | 0.96 (0.72, 1.28) | 0.78 | 1.00 (0.68, 1.50) | 0.98 |
| Hours of water delivery | | | | | | | |
| Continuous | 24 hrs | 3172 | 6.7 (211) | ref | -- | | |
| 1 | 7-24 hrs | 463 | 5.4 (25) | 0.84 (0.56, 1.26) | 0.40 | ref | -- |
| 2 | 5-6 hrs | 677 | 6.2 (42) | 0.87 (0.62, 1.23) | 0.43 | 1.04 (0.64, 1.69) | 0.87 |
| 4 | 4 hrs | 874 | 5.4 (47) | 0.85 (0.51, 1.40) | 0.51 | 1.01 (0.59, 1.75) | 0.97 |
| 5 | ≤3 hrs | 1164 | 9.5 (111) | **1.42 (1.08, 1.87)** | **0.01** | **1.69 (1.17, 2.44)** | **0.005** |
| Water availability (hours/day) | | | | | | | |
| Continuous | 24 hrs/day | 3172 | 6.5 (211) | ref | -- | | |
| 1 | 0.8-20 hrs/day | 623 | 6.4 (40) | 0.97 (0.58, 1.60) | 0.89 | ref | -- |
| 2 | 0.6-0.8 hrs/day | 602 | 7.3 (44) | 1.03 (0.73, 1.45) | 0.85 | 1.10 (0.64, 1.89) | 0.74 |
| 3 | 0.5-0.6 hrs/day | 377 | 6.1 (23) | 0.91 (0.47, 1.73) | 0.76 | 0.98 (0.42, 2.28) | 0.96 |
| 4 | 0.4-0.5 hrs/day | 900 | 5.9 (53) | 0.93 (0.58, 1.50) | 0.77 | 0.97 (0.55, 1.71) | 0.92 |
| 5 | <0.4 hrs/day | 676 | 9.6 (65) | **1.46 (1.04, 2.04)** | **0.03** | 1.50 (0.95, 2.34) | 0.08 |

aPR: Adjusted prevalence ratio; CI: Confidence interval
